# Characterization of the significant decline in humoral immune response six months post-SARS-CoV-2 mRNA vaccination: A systematic review

**DOI:** 10.1101/2021.12.10.21267593

**Authors:** Kin Israel Notarte, Israel Guerrero-Arguero, Jacqueline Veronica Velasco, Abbygail Therese Ver, Maria Helena Santos de Oliveira, Jesus Alfonso Catahay, Md. Siddiqur Rahman Khan, Adriel Pastrana, Grzegorz Juszczyk, Jordi B. Torrelles, Giuseppe Lippi, Luis Martinez-Sobrido, Brandon Michael Henry

**Affiliations:** Faculty of Medicine and Surgery, University of Santo Tomas, Manila, Philippines; Disease Intervention & Prevention and Population Health Programs, Texas Biomedical Research Institute, San Antonio, Texas, USA; Department of Biostatistics, State University of Maringa, Brazil; Department of Public Health, Medical University of Warsaw, Warsaw, Poland; Section of Clinical Biochemistry, University of Verona, Verona, Italy

**Keywords:** COVID-19, Pfizer-BioNTech, mRNA BNT162b2, Moderna, mRNA 1273, IgG, immunoglobulins, vaccines

## Abstract

Accumulating evidence shows a progressive decline in the efficacy of coronavirus disease 2019 (COVID-19) mRNA vaccines such as Pfizer-BioNTech (mRNA BNT161b2) and Moderna (mRNA-1273) in preventing breakthrough infections due to diminishing humoral immunity over time. Thus, this review characterizes the kinetics of anti-SARS-CoV-2 (Severe Acute Respiratory Syndrome Coronavirus 2) antibodies after the second dose of a primary cycle of COVID-19 mRNA vaccination. A systematic search of literature was performed and a total of 18 studies (*N*=15,980) were identified and reviewed. The percent difference of means of reported antibody titers were then calculated to determine the decline in humoral response after the peak levels post-vaccination. Findings revealed that the peak humoral response was reached at 21-28 days after the second dose, after which serum levels progressively diminished at 4-6 months post-vaccination. Additionally, results showed that regardless of age, sex, serostatus and presence of comorbidities, longitudinal data reporting antibody measurement exhibited a decline of both anti-receptor binding domain (RBD) IgG and anti-spike IgG, ranging from 94-95% at 90-180 days and 55-85% at 140-160 days, respectively, after the peak antibody response. This suggests that the rate of antibody decline may be independent of patient-related factors and peak antibody titers but mainly a function of time and antibody class/molecular target. Hence, this study highlights the necessity of more efficient vaccination strategies to provide booster administration in attenuating the effects of waning immunity, especially in the appearance of new variants of concerns (VoCs).

## Introduction

The coronavirus disease 2019 (COVID-19) pandemic claimed millions of lives worldwide and remains an unprecedented challenge to global public health.^1, 2^ Despite the increasing availability of therapeutics against Severe Acute Respiratory Syndrome Coronavirus 2 (SARS-CoV-2) infection, mass vaccination is the mainstay for limiting new infections, reinfections, breakthrough infections, and unwanted sequelae from COVID-19.^3–6^ Although nationwide immunization programs have already been implemented in most countries, the mass production, allocation, and accessibility to COVID-19 vaccines remain a hurdle, especially in low-income countries where administration of primary and adjunctive doses could be challenging.^7^

Among different types of COVID-19 vaccines, the utilization of mRNA-based vaccines is unprecedented.^8^ Despite the efficacy of mRNA vaccines in reducing the risk of developing severe COVID-19 illness, ample evidence showed the progressive decline in their efficacy for preventing SARS-CoV-2 infections.^9^ Such vulnerability is correlated to the significant decline in anti-SARS-CoV-2 antibodies (Abs) over time.^10–12^ This waning humoral response could lead to breakthrough infections, thus highlighting the potential role of serologic testing to assess vaccine immunogenicity and protection efficacy. While the role of T cells and B memory cells from vaccine-induced immunity has yet to be fully elucidated, multiple studies showed that Ab levels directly correlate with risk of vaccine reinfection and breakthrough, with low levels of anti-SARS-CoV-2 IgG in patients with infections correlating with high viral load and duration of viral shedding.^3, 4^ In addition, the emergence of the new SARS-CoV-2 Omicron variant (B.1.1.529), with potential for high immune escape, highlights the necessity for vaccine boosters and high Ab titers for an improved immunoprotection.^13^ Thus, to better understand the rate of decay of anti-SARS-CoV-2 Abs through time, this rapid systematic review was designed to analyze humoral response within a 6**-**month period among recipients of the Pfizer-BioNTech or Moderna COVID-19 mRNA-based vaccines to provide further evidence on the necessity of boosting vulnerable populations.

## Methods

### Search strategy and eligibility criteria

A systematic literature search was conducted to identify studies reporting the decline of humoral response of individuals post-mRNA vaccination. As shown in **Figure 1**, a comprehensive search was carried out in PubMed, Cochrane CENTRAL, Google Scholar, Science Direct, medRxiv, and Research Square for articles published from January 2021 to December 2021. The search keywords used included “SARS-CoV-2”, “COVID-19”, “declining humoral response”, “post-vaccination”, “mRNA vaccine”, “Pfizer-BioNTech”, “Moderna”, “mRNA-1273”, and “mRNA BNT162b2” which resulted in 51 journal articles.

**Figure 1.**
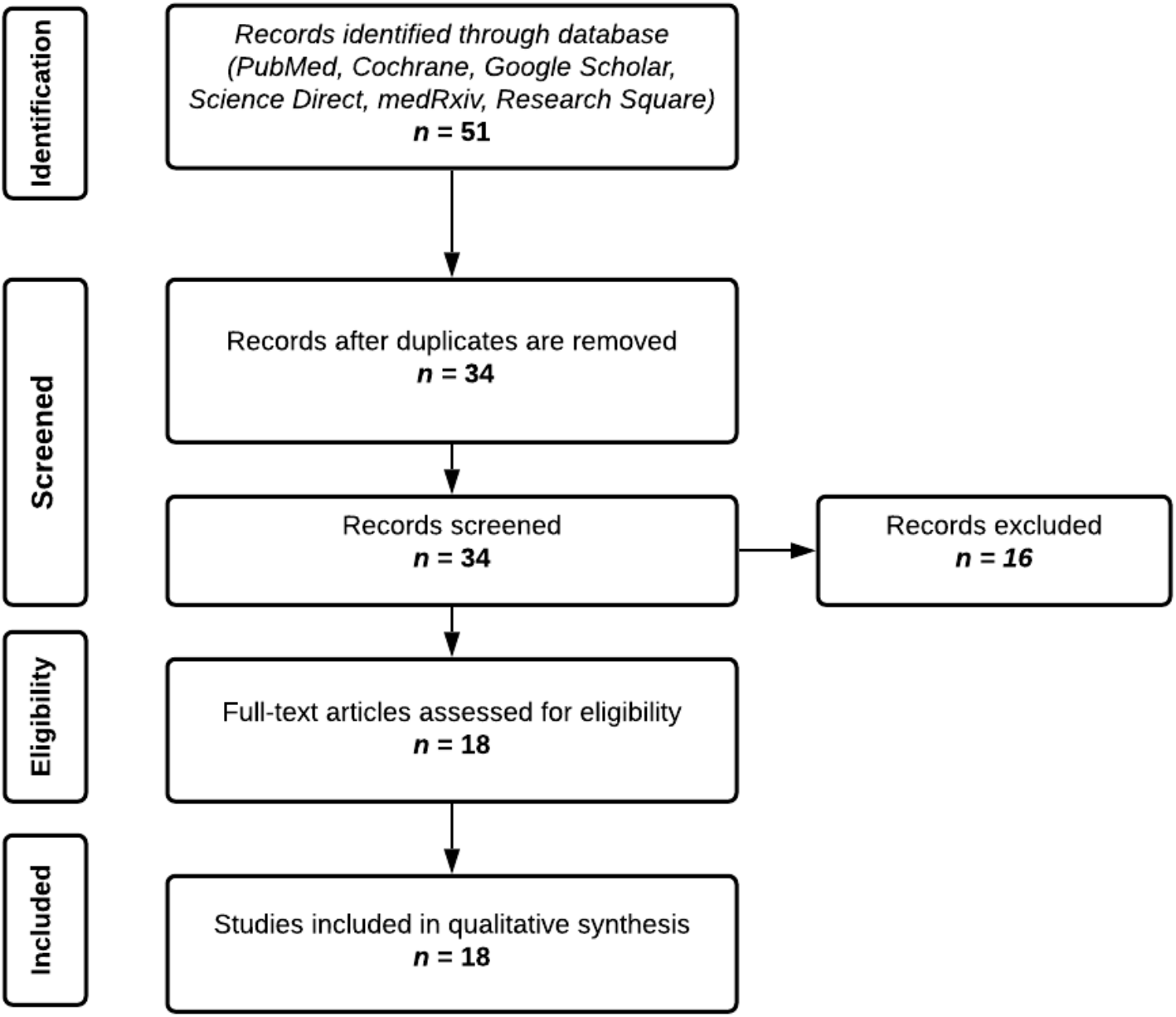
Screening and appraisal of journal articles for inclusion in the systematic review.

For the inclusion criteria, articles reporting the following data were considered: (1) all participants who received a primary vaccination cycle with two complete doses of Pfizer-BioNTech (mRNA BNT162b2) or Moderna (mRNA-1273), (2) with at least a record of 10 patients and above (3) individual IgG or IgA, total Ig, or neutralizing anti-SARS-CoV-2 antibody titers, (4) quantitative or semi-quantitative antibody tests, (6) articles reporting humoral response 4-8**–** months post-vaccination, (7) articles available in the English language, and (8) randomized controlled, cohort, preprint or published papers and (9) providing complete and extractable data given the limited papers available for this novel disease and the mRNA vaccine.

The exclusion criteria were: (1) participants who received a single dose of mRNA vaccine only, (2) participants who received other types of vaccines such as Sinovac, AstraZeneca, Johnson & Johnson, Novavax, and Sanofi-GSK, (3) articles reporting humoral response <4 months post-vaccination, and (4) anti-SARS-CoV-2 IgM response and cell-mediated immunity; the former because it is now increasingly clear that IgM plays a minor role against COVID-19, as it has lower sensitivity (64%), specificity (99%), and accuracy (94%) compared to IgG (93%, 100%, and 98%, respectively).^14^ IgM antibodies were also found to decline early (*i.e*., at day 20 post-vaccination) and have lower neutralizing potential.^15^ The latter because we only focused on humoral immunity due to limited data and lack of standardization of available cell-mediated immunity assays, hence imposing difficulty in data analysis.

### Data extraction

Data were extracted independently from each article by two authors into a spreadsheet. A third author checked the extracted data for completeness and accuracy. Any disagreements were resolved by consensus among the authors.

Descriptive and outcome data were extracted from the included studies, including type of vaccine, country of origin, sample size, age range or median age of the population, type of serologic test employed and its manufacturing company, analyzer used, immunoglobulin measured, and molecular target of immunoglobulin. In addition, serial or non-serial serologic measurement determination was done, to differentiate which study performed better patient follow-up, thus reducing methodologic bias due to greater accountability for inter-individual variability despite general lower sample size for serial serologic sampling. Additional data were requested from the original study authors when necessary.

### Data analysis

All data reported in this study were in reference to the peak humoral response after a primary vaccination cycle with 2 doses. However, due to significant heterogeneity in the assays used to probe antibody titers, the antibody measurements reported in the articles were standardized using the percent difference of means. This shows the absolute value of the ratio of the difference between two groups, (groups A and B which pertain to antibody titer measurements on the peak and after the peak of humoral response) and their average, expressed as a percentage to enable standard comparison of these data regardless of their units of measurements and the diagnostic tools used for their quantification. It is computed using the formula below:

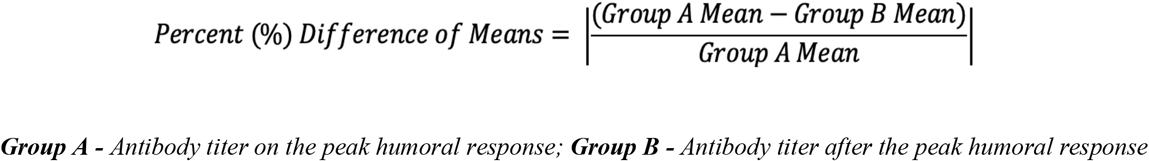

Therefore, values are reported as percentual decrease from the peak. Time points were also standardized to represent number of days after the second vaccine dose.

### Scope and limitations

The demographic parameters used in this study were limited to age, sex, serostatus, and comorbidities such as hemodialysis or chronic kidney disease, immunologic disorders, metabolic derangements including heart disease, hypertension, and diabetes mellitus. It also includes a general section wherein no specific parameter was identified in determining the percent decrease in humoral response. These factors were analyzed independently of each other.

Moreover, the authors utilized both preprint and published journal articles written from January 2021 up to the present which included vaccination time points from days 0 to 201 of either Pfizer-BioNTech (mRNA BNT162b2) or Moderna (mRNA-1273) in demonstrating the decline of humoral response. These vaccines were only discussed in this systematic review as they were the leading vaccine utilized worldwide and due to the wide range of available resources at the time of writing.

A quantitative meta-analysis was not done because of the heterogeneity determined between studies, as well as between different immunoassays used in each study, their units, sensitivities/specificities, and antigenic targets, not appropriate to perform a reliable and sensible pooled analysis. Thus, percent (%) differences in titers between groups were calculated, as an attempt to standardize the antibody data and enable a combined evaluation of different studies.

Lastly, we did not evaluate cell-mediated immunity and the kinetics of T cells after vaccination. The measurement of T-cell responses against SARS-CoV-2 is complex and lacks standardization to allow for comparisons in the literature, but is a priority topic for future research. Nonetheless, Gilbert et al. demonstrated that the majority (68.5%) of vaccine efficacy is mediated by neutralizing antibody responses, with the remaining 31.5% of vaccine efficacy may be attributed to cell-mediated responses, other functional antibodies, and innate immunity.^16^ These findings are in agreement with studies demonstrating the relationship between neutralizing antibody titers and breakthrough infections.^3, 4^

## Results

A total of 18 articles (*N*=15,980) were included in this systematic review in which 14 studies focused on Pfizer-BioNTech (*n*=13,364), 2 on Moderna (*n*=234), and 2 included both Pfizer-BioNTech and Moderna (*n*=2,382). These were further divided into four categories, humoral immunity post-second dose vaccination influenced by age, sex, baseline serostatus, and presence of comorbidities. Thirteen articles were included in the general section, four under the age category, four under sex, four under serostatus, and three articles under comorbidities. For the sex category, a total of 3,128 males and 6,237 females were included in the analysis. Meanwhile, the comorbidities cohort were further grouped according to the reported condition. Three studies reported on hemodialysis or end stage renal disease, one study reported on heart disease and hypertension, one for diabetes mellitus, and one for immunologic disorders. It is also worth noting that articles under these categories were non-exclusive, as most studies included more than one factor in their analysis. With respect to the geographical distribution, seven studies were from Italy, three studies from the United States, three from Israel, two from Belgium, and one each from Japan, Estonia and Austria. Of the reviewed articles that conducted serial study sampling, the largest sample size was from Italy (*n*=787).

**Tables 1–5** provide a summary of the following trends observed in this study. In general, there is a substantial decrease of Ab titers against SARS-CoV-2 following 6 months post-second dose mRNA vaccination (**Figure 2**); however, the amount and rate of decline of the Ab titers varied according to the factors analyzed. Participants exhibited up to 95% decline in Ab levels at least 120 days after the peak rise of the IgG titers post-mRNA vaccination. In studies reporting longitudinal data on Ab measures, an observed decline from peak levels of anti-RBD IgG ranged from 94 to 95% at 90 to 180 days post-peak, while the anti-spike IgG showed a decline that ranged from 55 to 85% at 140 to 160 days after the peak (**Figure 2**). With respect to the vaccine brand, Pfizer-BioNTech displayed a 90 to 94% decline of anti-RBD IgG at 150 to 161 days following the peak, and a decline of IgG anti-spike protein ranging from 55 to 95% at 140 to 180 days after the peak. Meanwhile, Moderna exhibited a 69 to 96% decrease in anti-RBD IgG at 150-174 days from the peak, and a 45% decline in anti-spike IgG 90 days post-titer peak.

**Table 1.**
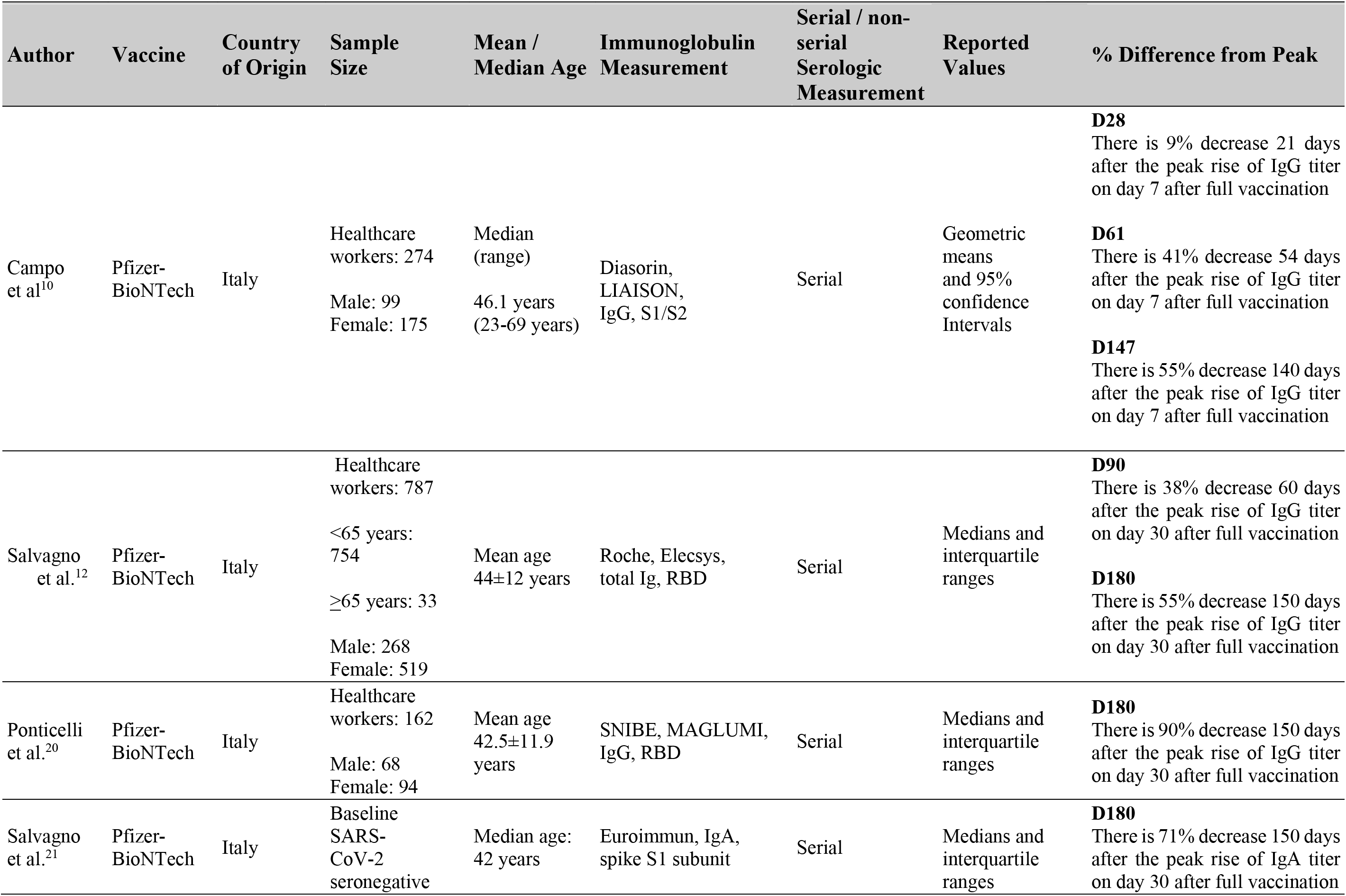
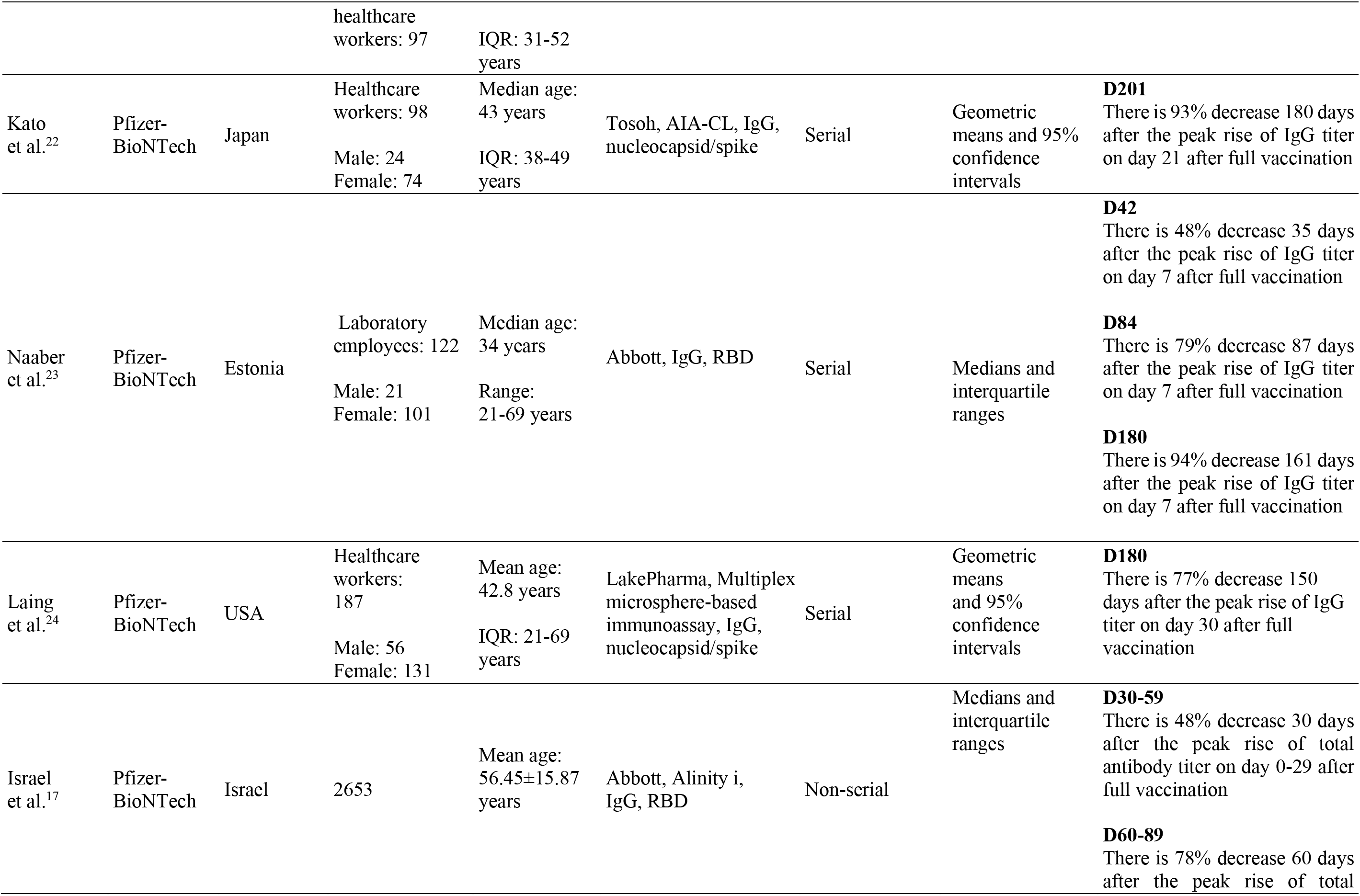
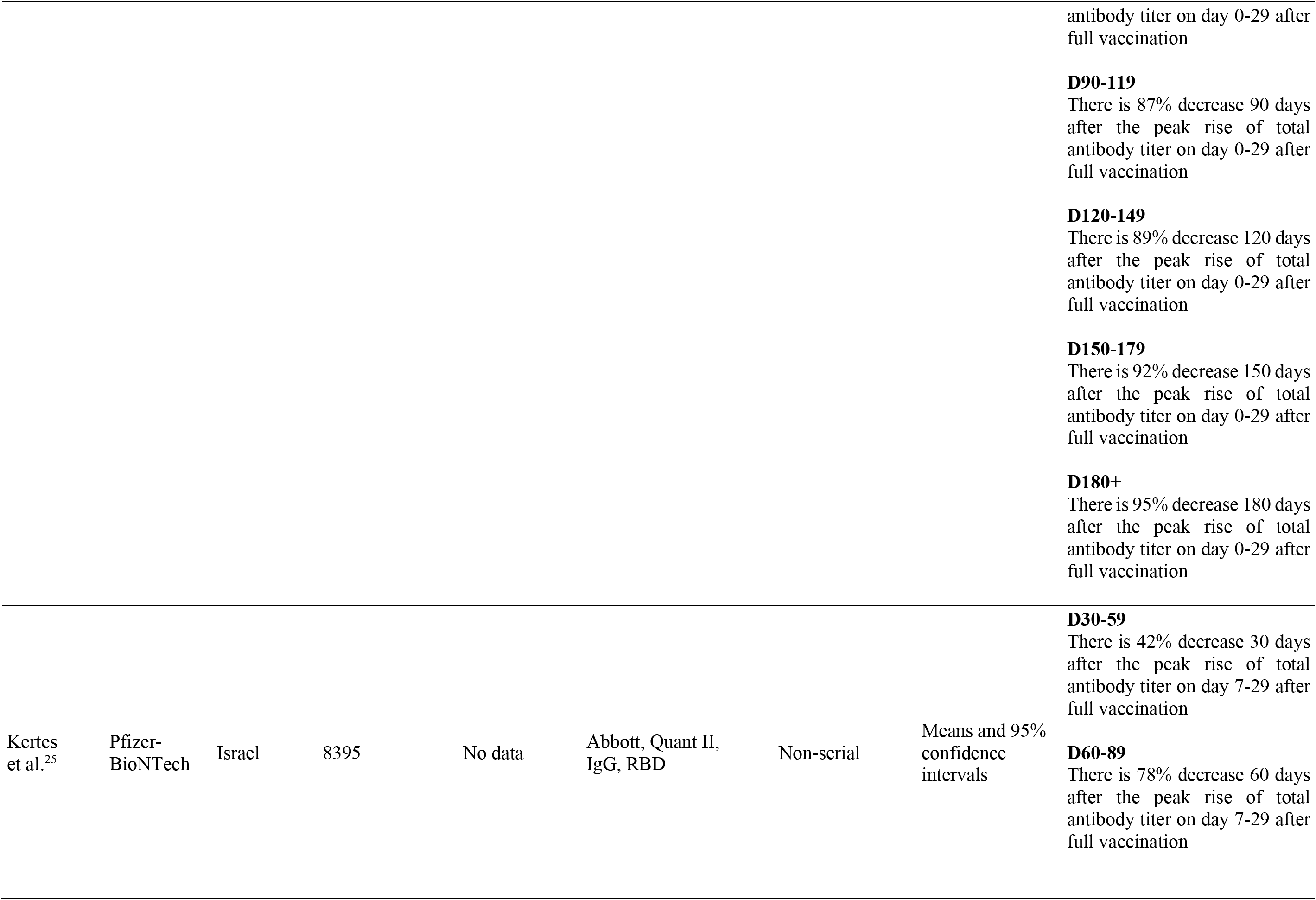
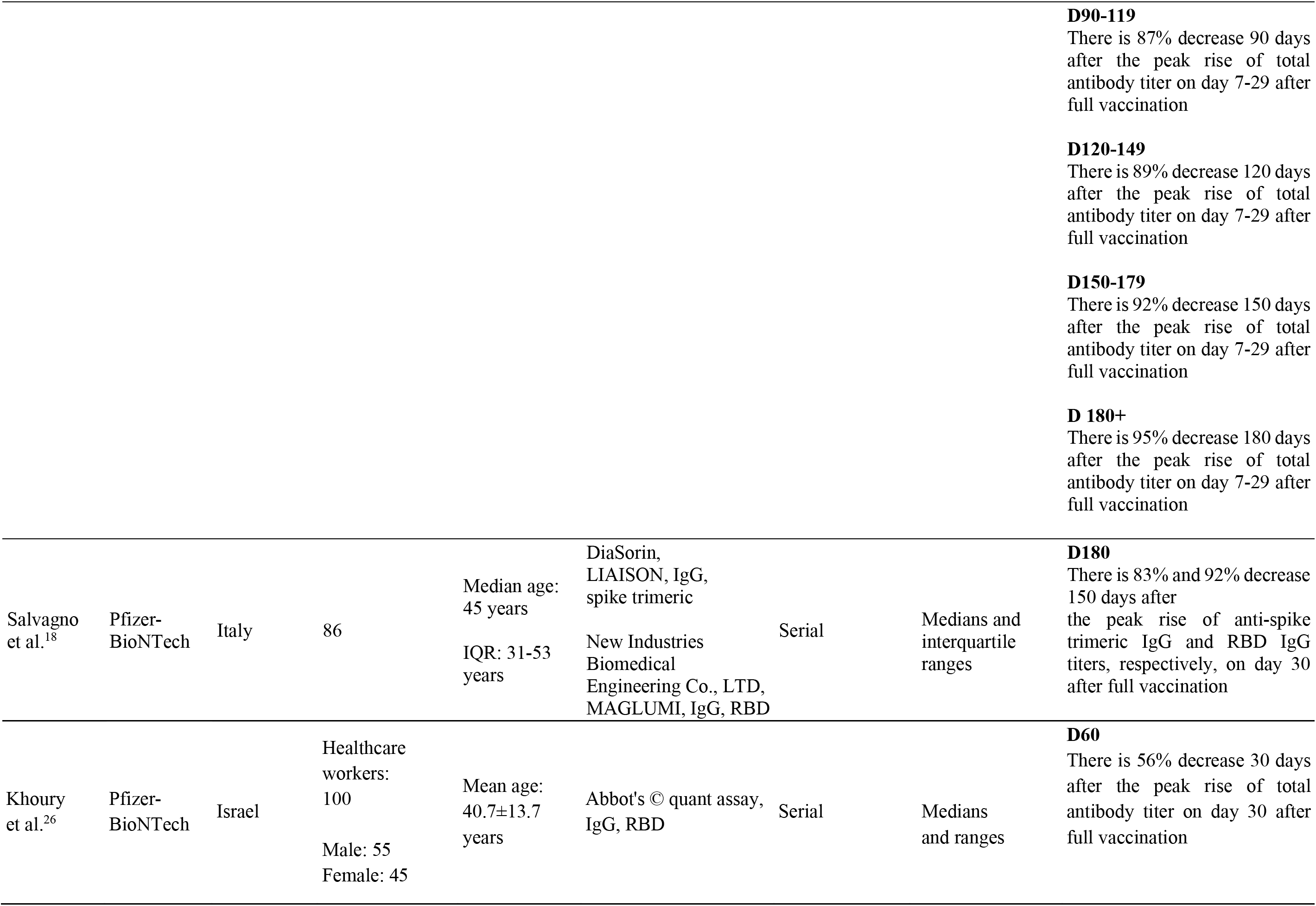
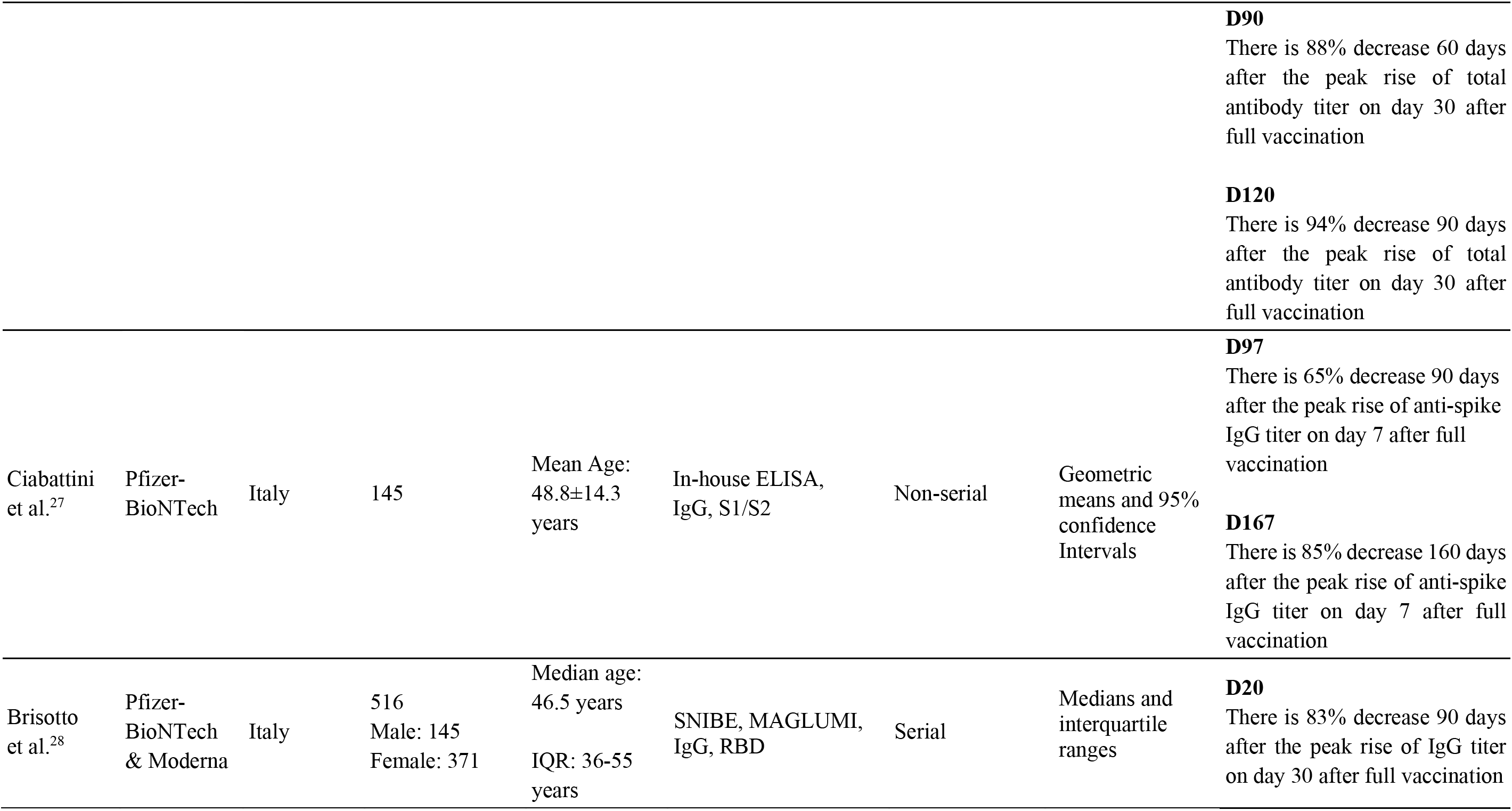
Declining humoral response post-second dose administration of SARS-CoV-2 mRNA vaccine.

**Table 2.**
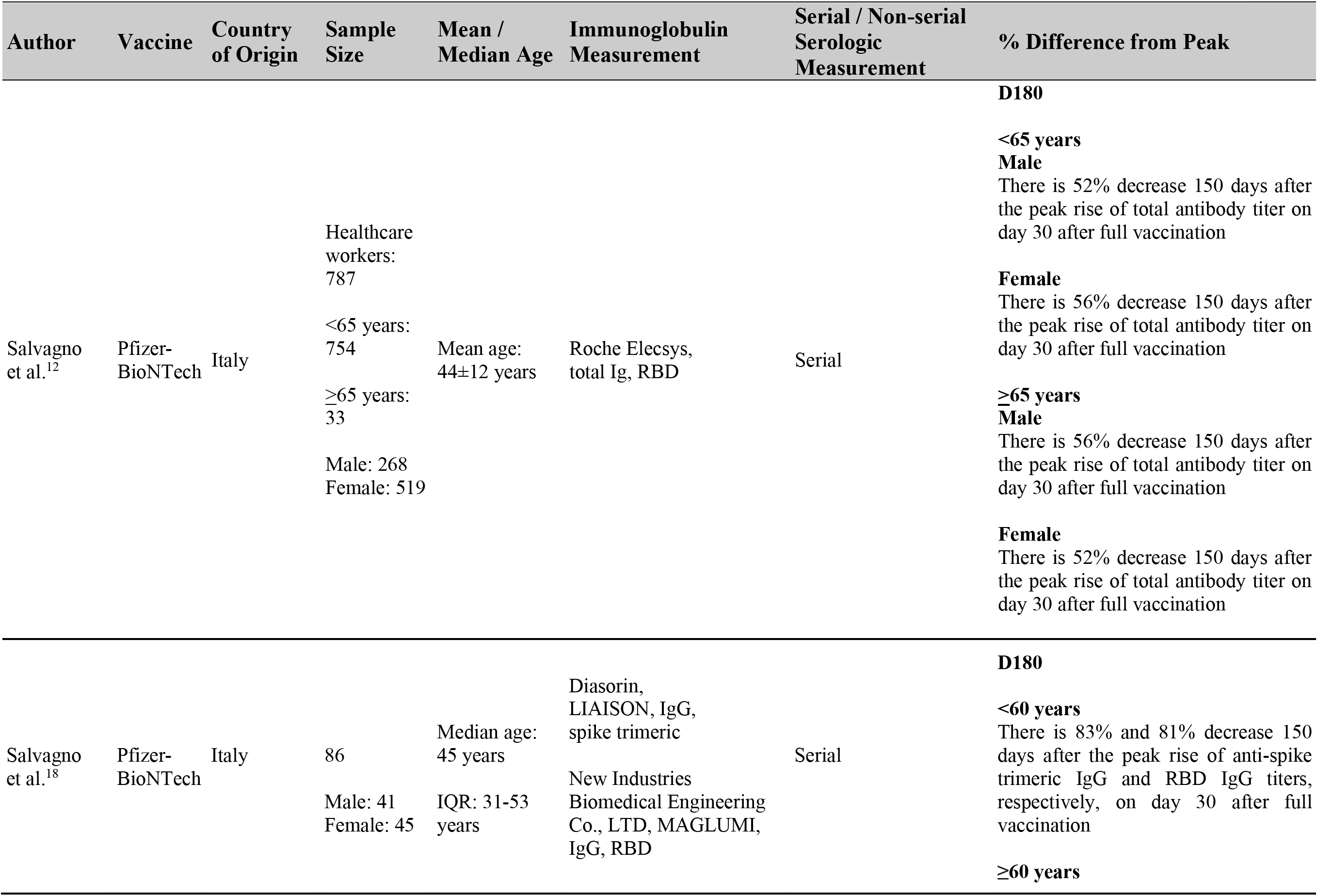
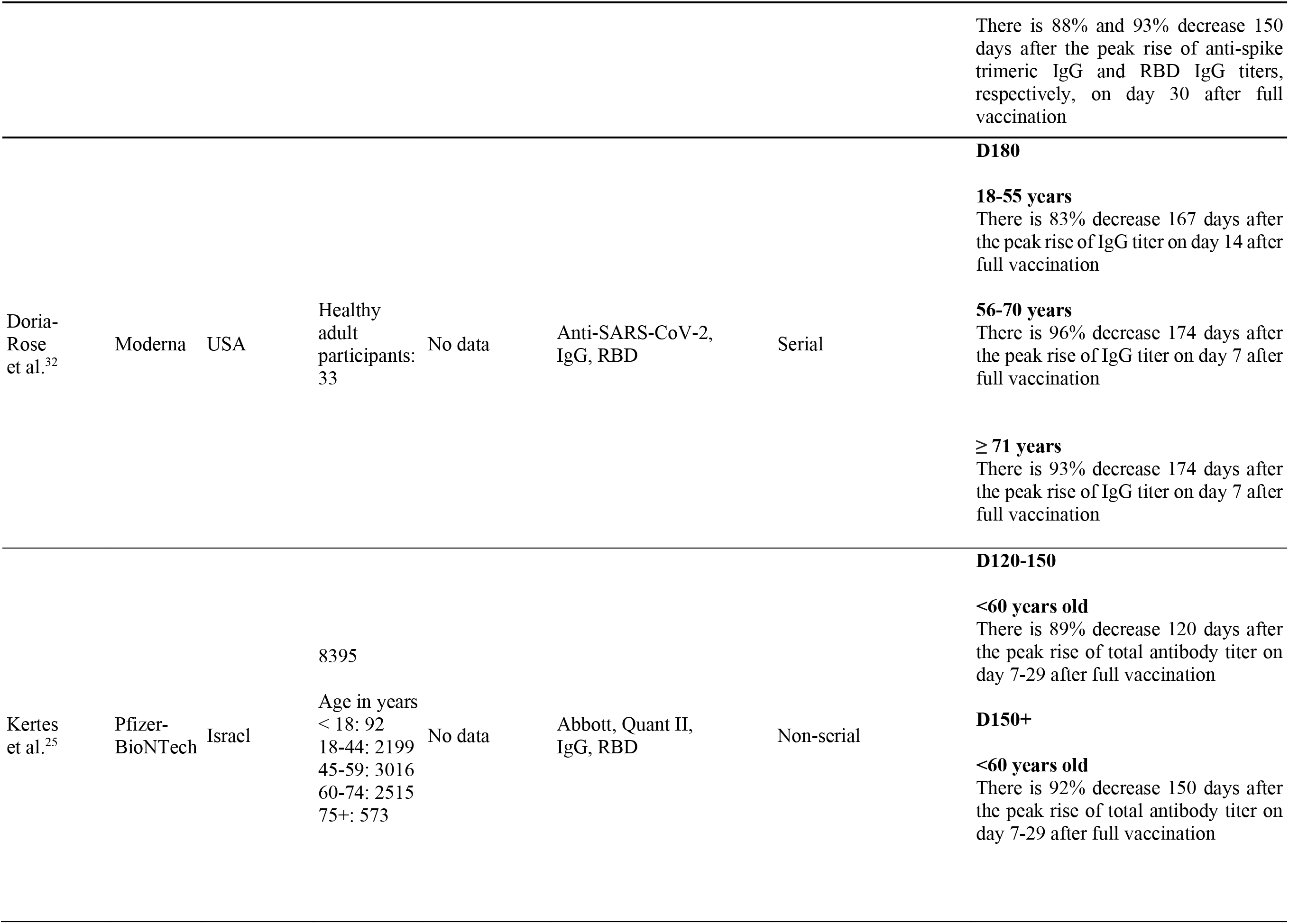
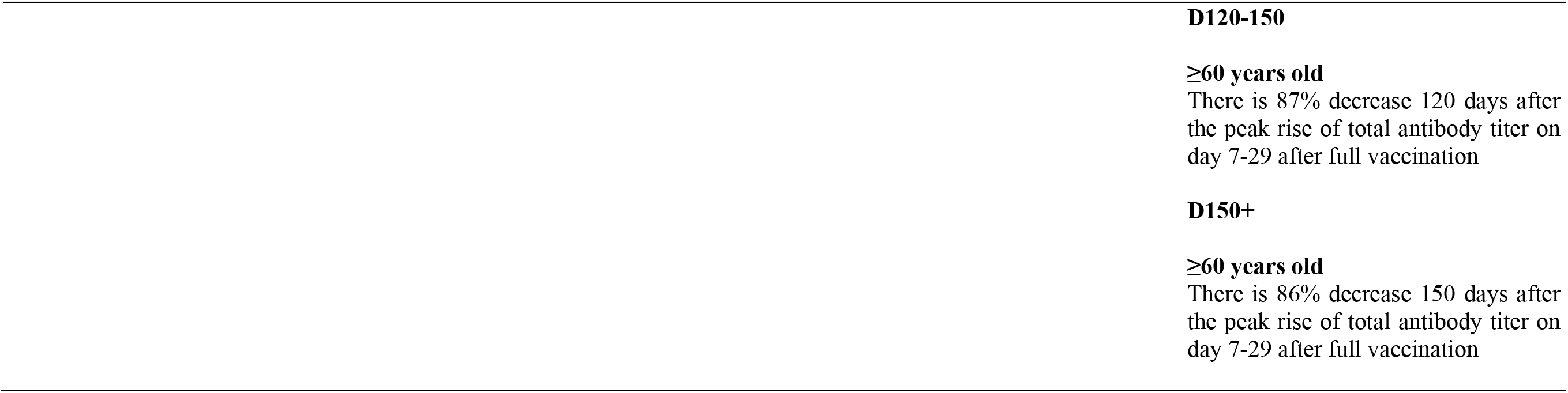
Effect of age in declining humoral response post-second dose administration of SARS-CoV-2 mRNA vaccine.

**Table 3.**
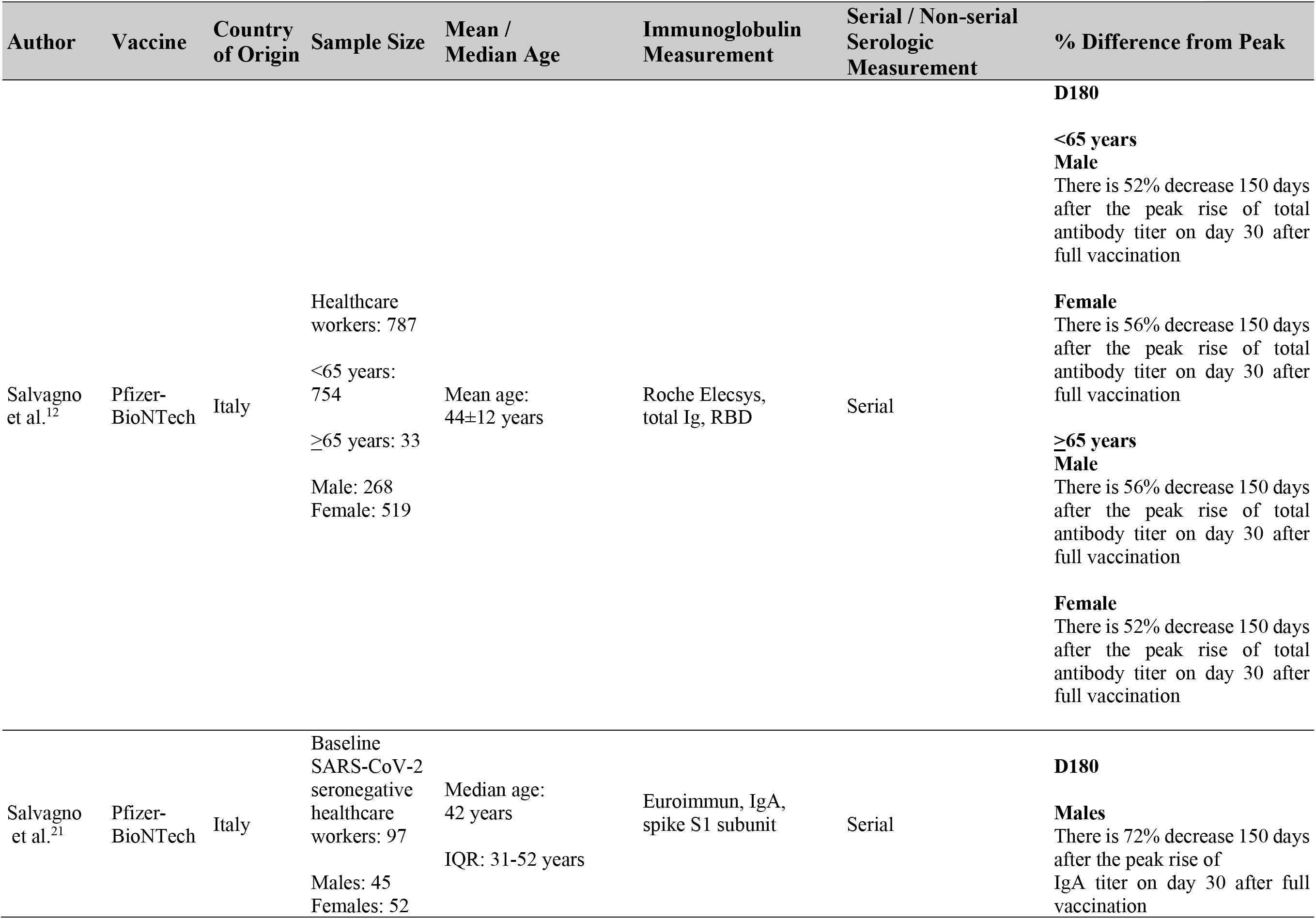
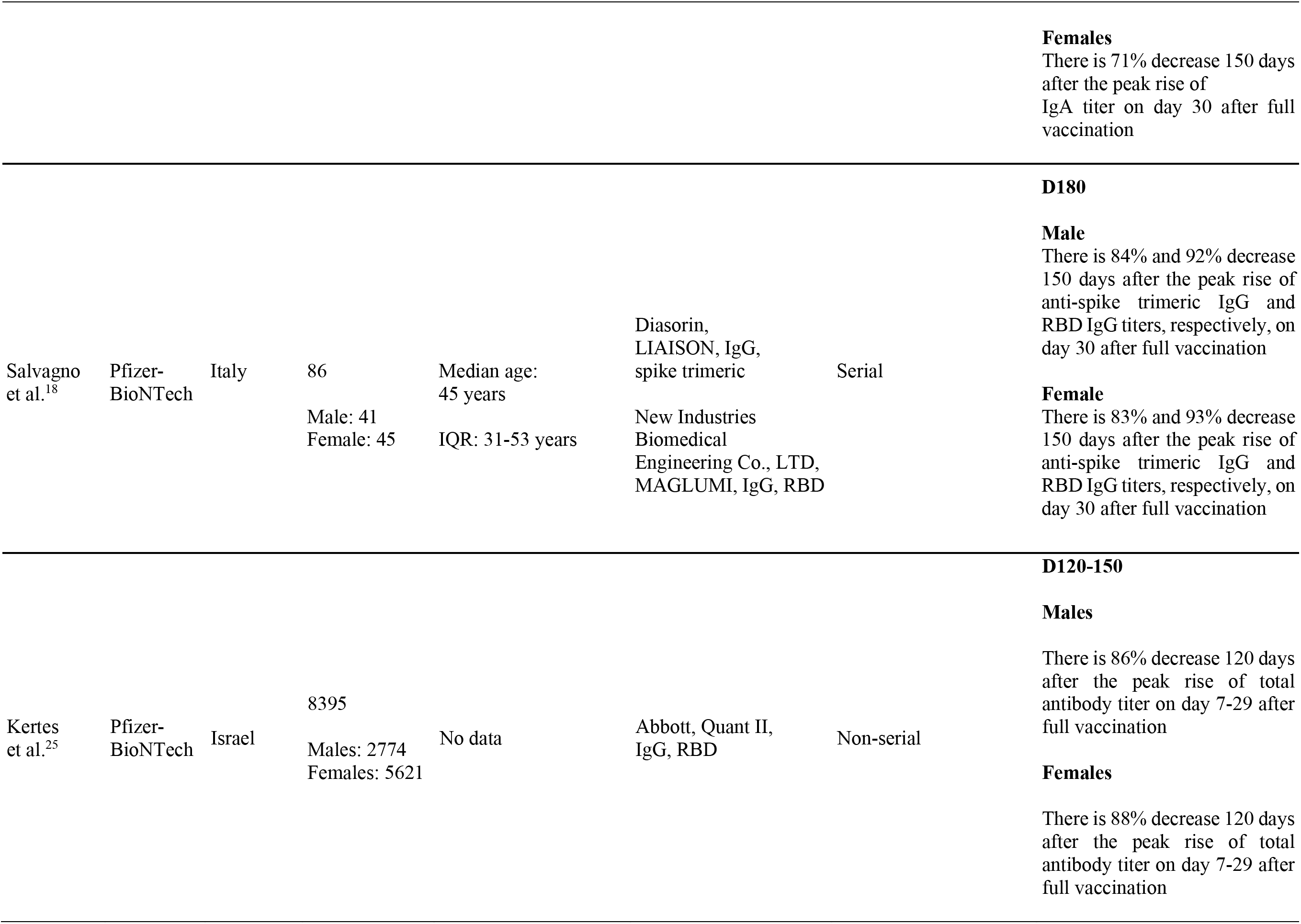
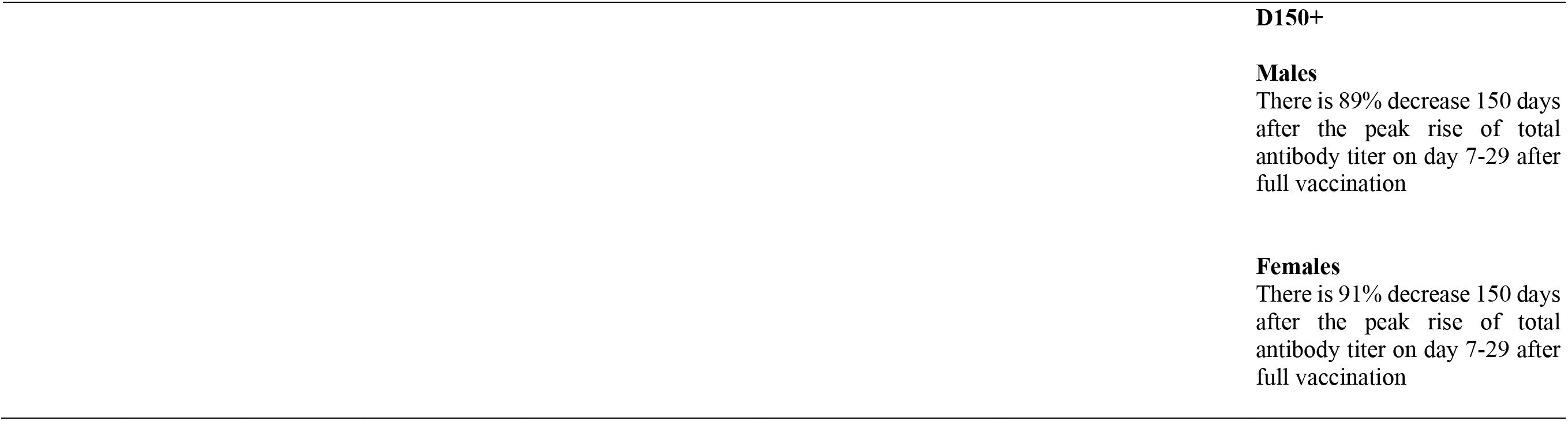
Effect of sex in declining humoral response post-second dose administration of SARS-CoV-2 mRNA vaccine.

**Table 4.**
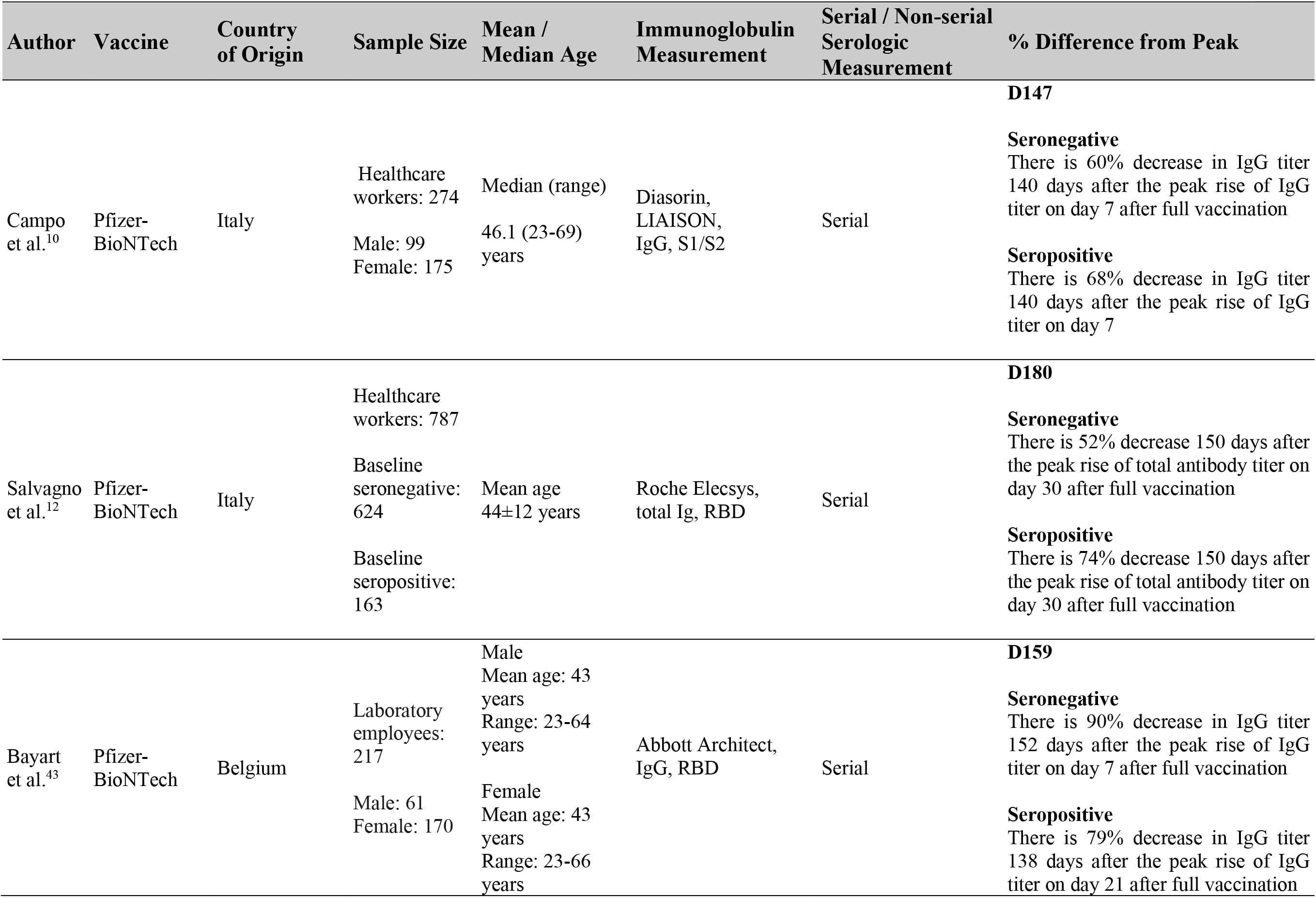
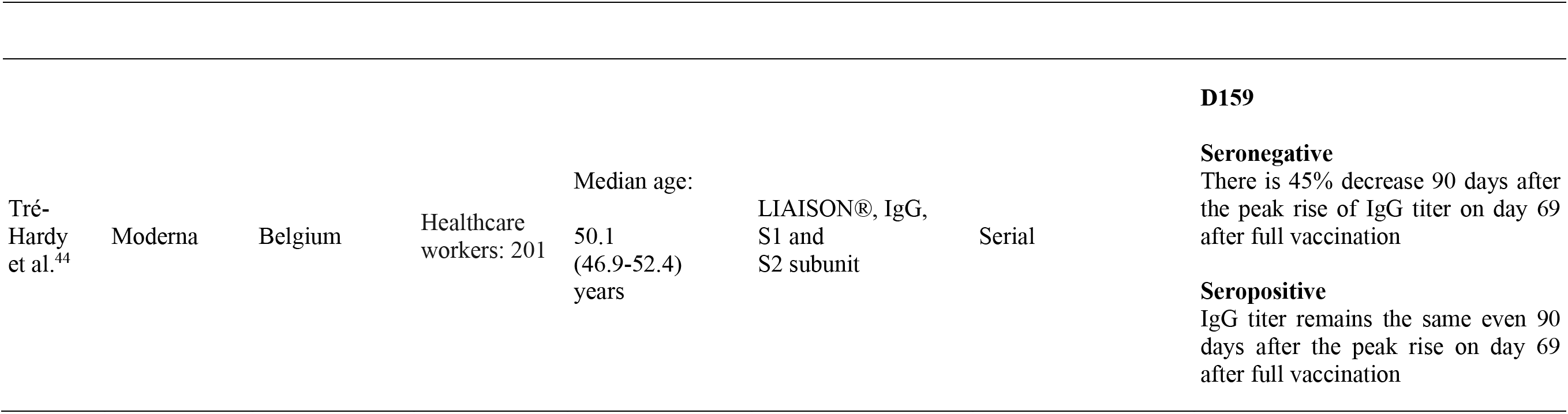
Effect of serostatus in declining humoral response post-second dose administration of SARS-CoV-2 mRNA vaccine.

| Author | Vaccine | Country of Origin | Sample Size | Mean / Median Age | Immunoglobulin Measurement | Serial / Non-serial Serologic Measurement | % Difference from Peak |
| --- | --- | --- | --- | --- | --- | --- | --- |
|  |  |  |  |  |  |  | **D147** |
| Campo et al.<sup>10</sup> | Pfizer-BioNTech | Italy | Healthcare workers: 274<br>Male: 99<br>Female: 175 | Median (range)<br>46.1 (23-69) years | Diasorin, LIAISON, IgG, S1/S2 | Serial | **Seronegative**<br>There is 60% decrease in IgG titer 140 days after the peak rise of IgG titer on day 7 after full vaccination<br>**Seropositive**<br>There is 68% decrease in IgG titer 140 days after the peak rise of IgG titer on day 7 |
|  |  |  |  |  |  |  | **D180** |
| Salvagno et al.<sup>12</sup> | Pfizer-BioNTech | Italy | Healthcare workers: 787<br>Baseline seronegative: 624<br>Baseline seropositive: 163 | Mean age 44±12 years | Roche Elecsys, total Ig, RBD | Serial | **Seronegative**<br>There is 52% decrease 150 days after the peak rise of total antibody titer on day 30 after full vaccination<br>**Seropositive**<br>There is 74% decrease 150 days after the peak rise of total antibody titer on day 30 after full vaccination |
|  |  |  |  |  |  |  | **D159** |
| Bayart et al.<sup>43</sup> | Pfizer-BioNTech | Belgium | Laboratory employees: 217<br>Male: 61<br>Female: 170 | Male<br>Mean age: 43 years<br>Range: 23-64 years<br>Female<br>Mean age: 43 years<br>Range: 23-66 years | Abbott Architect, IgG, RBD | Serial | **Seronegative**<br>There is 90% decrease in IgG titer 152 days after the peak rise of IgG titer on day 7 after full vaccination<br>**Seropositive**<br>There is 79% decrease in IgG titer 138 days after the peak rise of IgG titer on day 21 after full vaccination |
|  |  |  |  |  |  |  | **D159** |
| Tré-Hardy et al.<sup>44</sup> | Moderna | Belgium | Healthcare workers: 201 | Median age: | LIAISON®, IgG, S1 and S2 subunit | Serial | **Seronegative**<br>There is 45% decrease 90 days after the peak rise of IgG titer on day 69 after full vaccination |
|  |  |  |  | 50.1 (46.9-52.4) years |  |  | **Seropositive**<br>IgG titer remains the same even 90 days after the peak rise on day 69 after full vaccination |

**Table 5.**
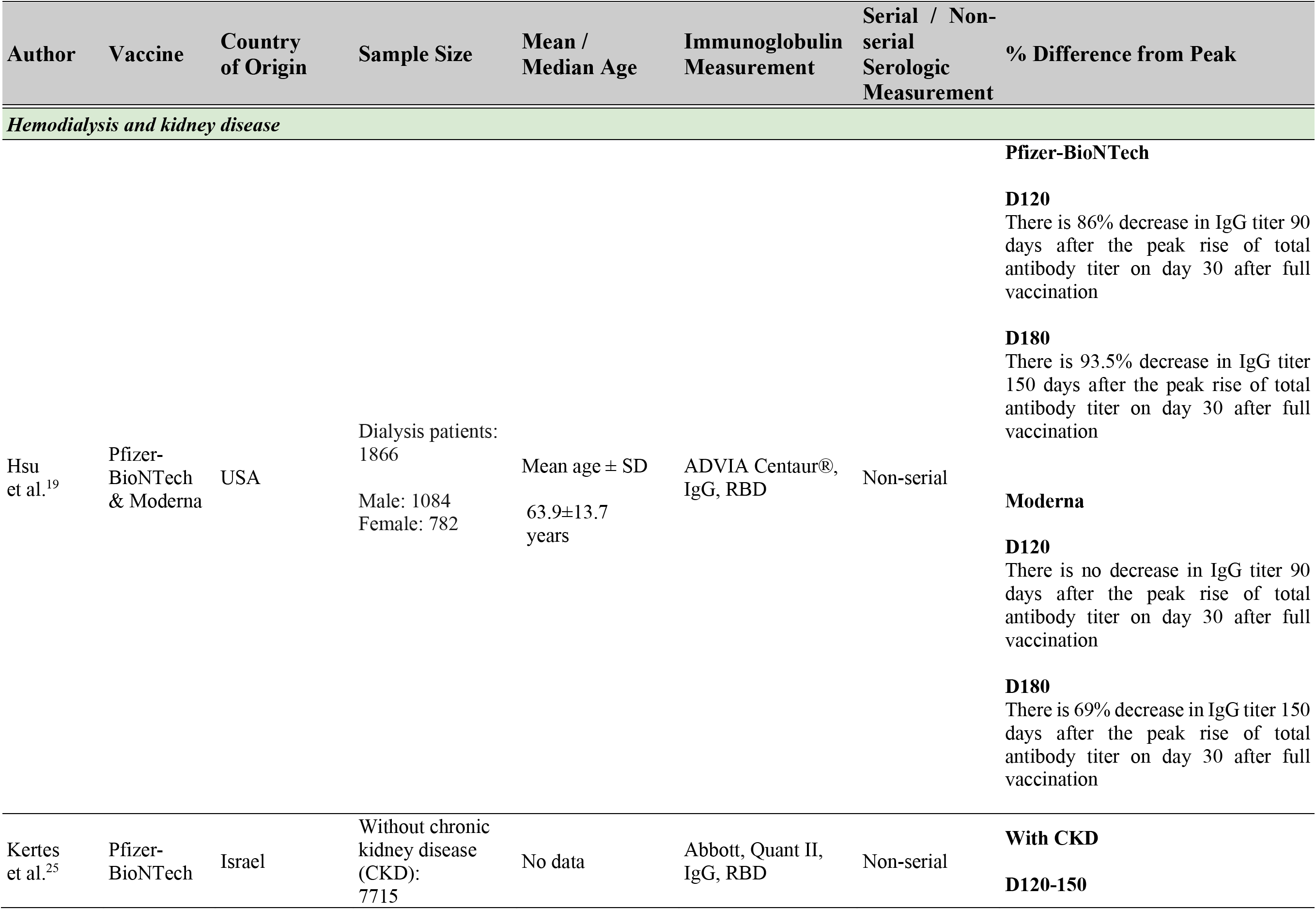
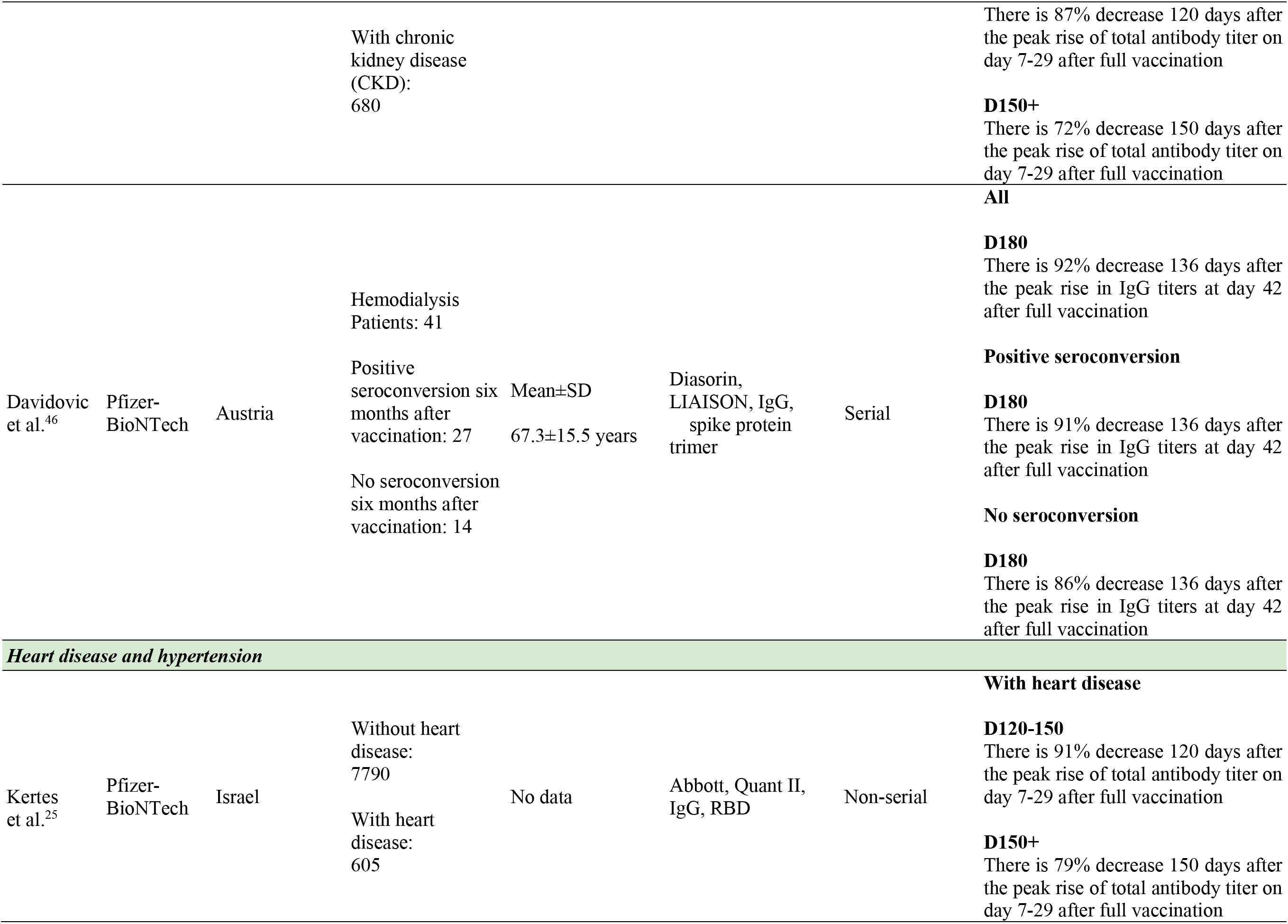
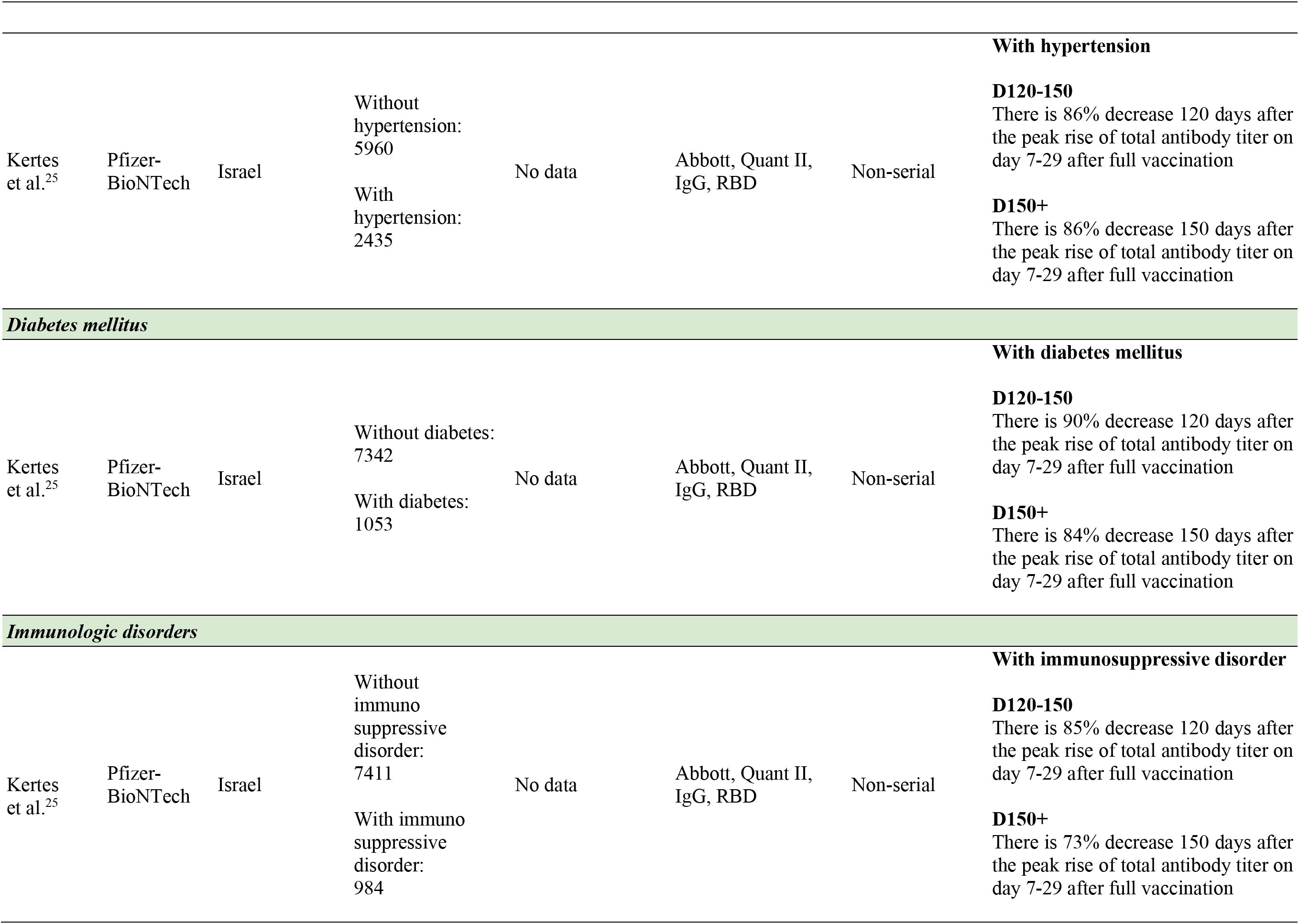
Effect of comorbidities in declining humoral response post-second dose administration of SARS-CoV-2 mRNA vaccine.

**Figure 2.**
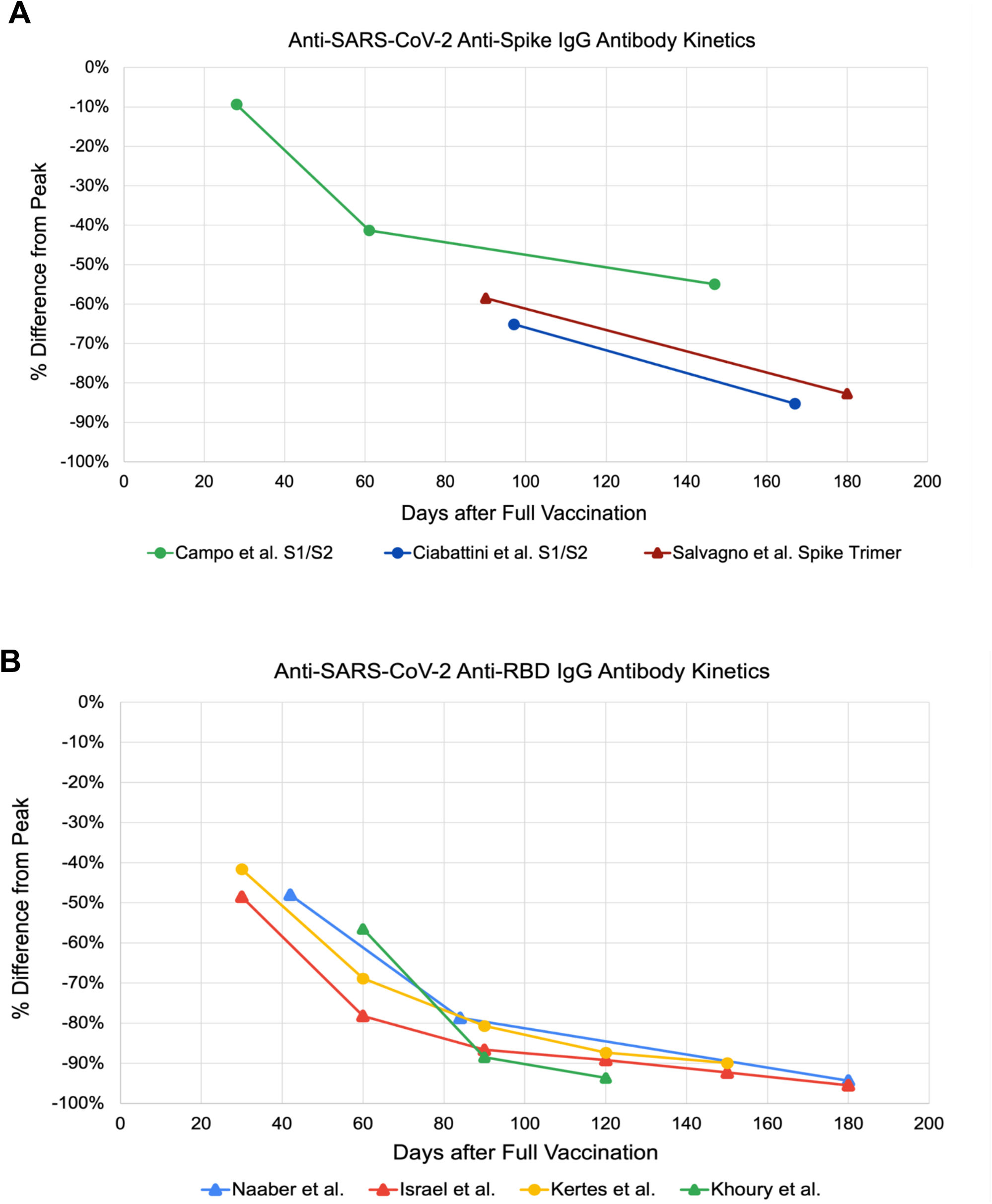
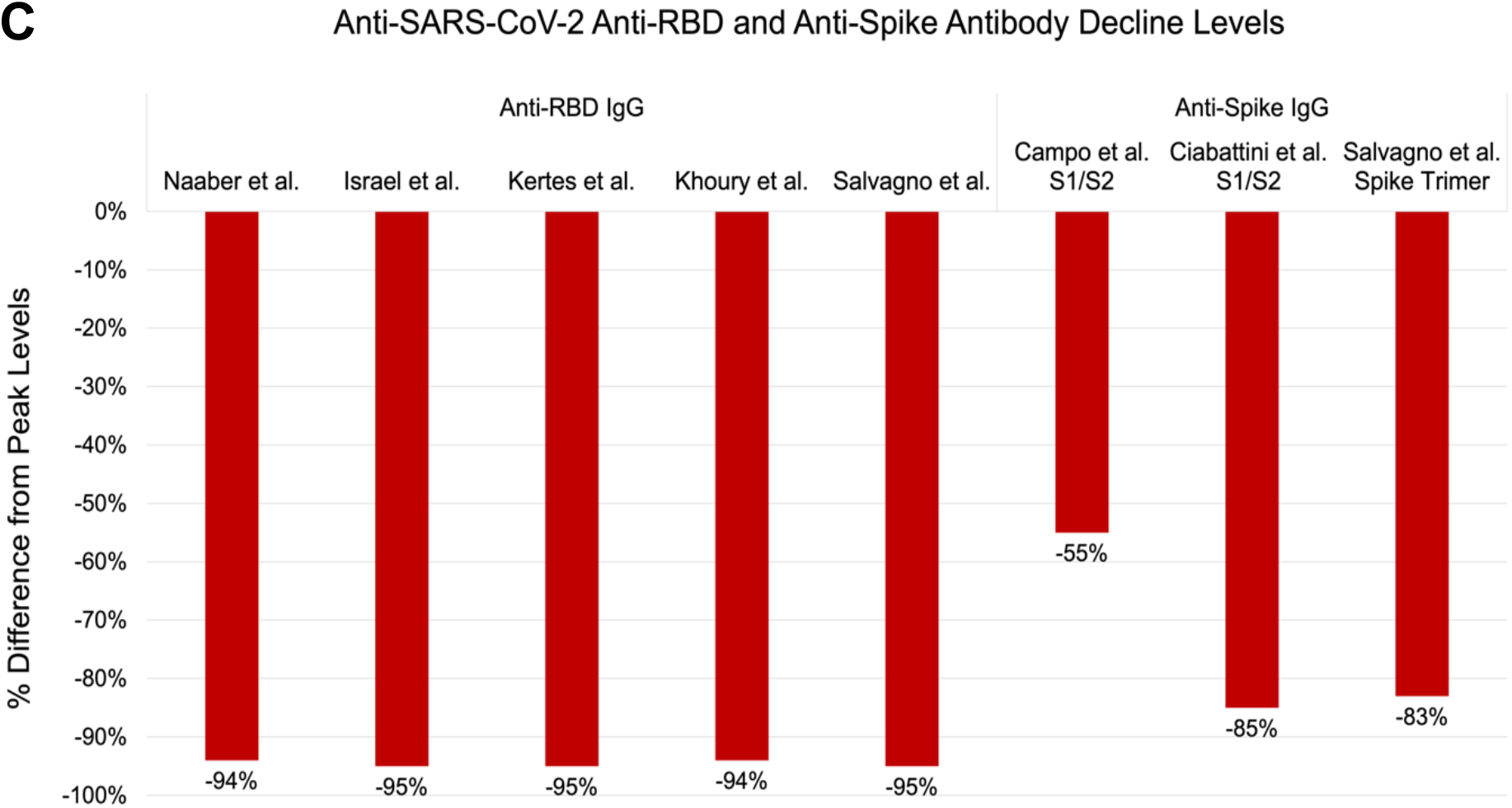
Kinetics of antibodies in studies reporting longitudinal data showing percentage decline from peak titer after vaccination for anti-spike **(A)** and anti-RBD **(B)** IgG antibodies. Data summary of antibody percentage decline levels at ∼6 months post-vaccination **(C)**.

A consistent decrease in humoral immunity post-vaccination was found across all age groups ranging from 53 to 96% (**Figure 3**). Similarly, a consistent decline in Ab titers regardless of sex was also noted. Females reported a 91 to 93% decrease in their anti-RBD IgG levels, similar to males (ranging from 89 to 92%) (**Figure 4**). Anti-spike IgG levels in females decreased by 83%, also close to the decline seen in males (84%) (**Figure 4**). Regardless of serostatus, a diminished humoral immunity post-mRNA vaccination was evident. However, the Ab titers post-second dose of mRNA vaccine displayed varying trends (**Figure 5**). One study showed a 90% decrease of titers in the seronegative group, which is higher than the decline observed in the seropositive group (79%) (**Figure 5**). This is in contrast with two studies, one showing a 52% and the other a 60% decline in their seronegative participants, which is lower than the observed decline in their seropositive participants (68% and 74%, respectively) (**Figure 5**). For comorbidities cohorts, there was a consistent decrease in the total antibody and IgG levels among those individuals with kidney disease and undergoing hemodialysis (93%), heart disease and hypertension (91%), diabetes mellitus (90%), and immunologic disorders (85%).

**Figure 3.**
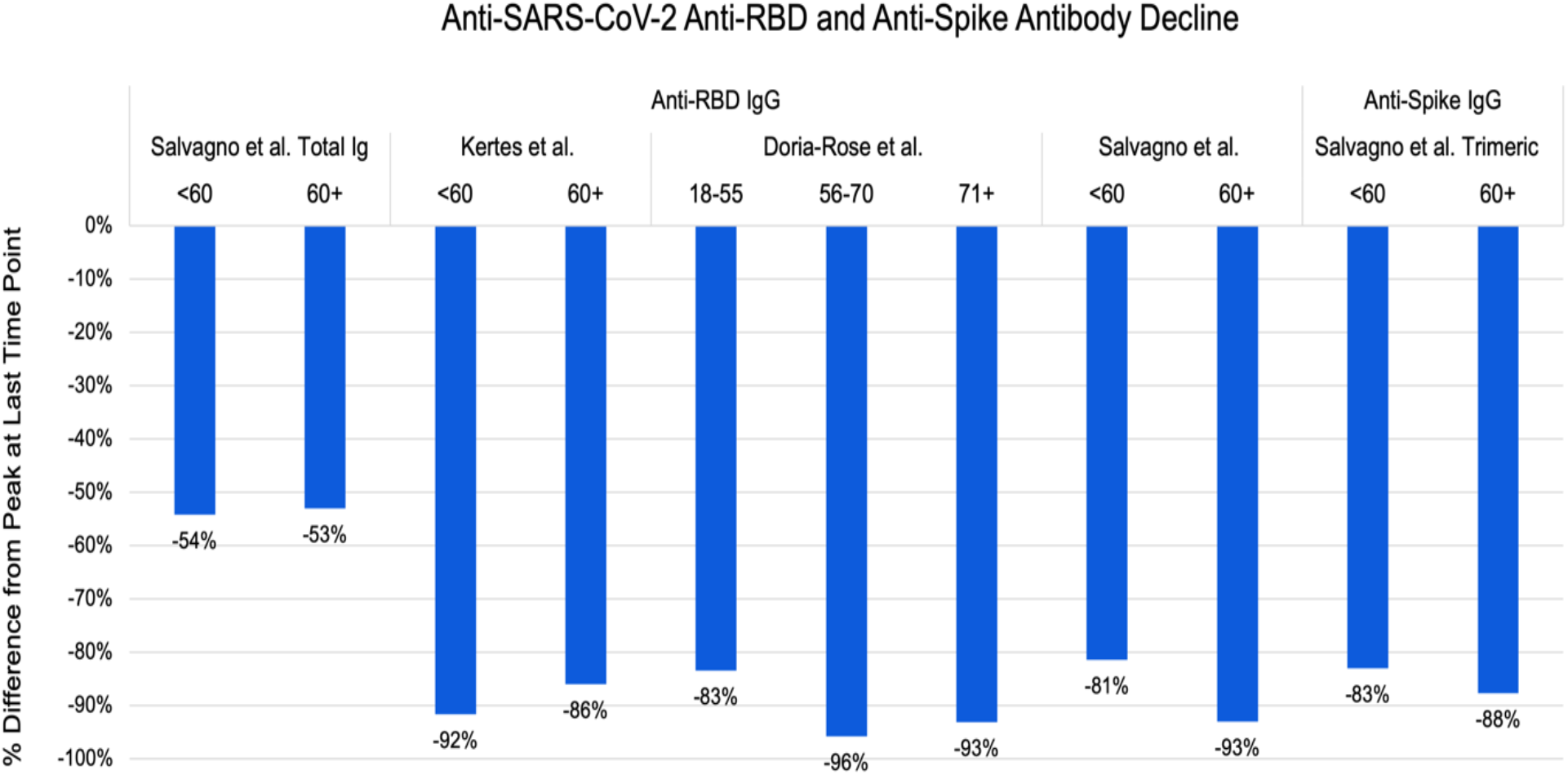
Comparison by age of the percentage decline of anti-SARS-CoV-2 antibodies from peak to last measured timepoint (∼ 6 months) in each study by immunoglobulin class and molecular target.

**Figure 4.**
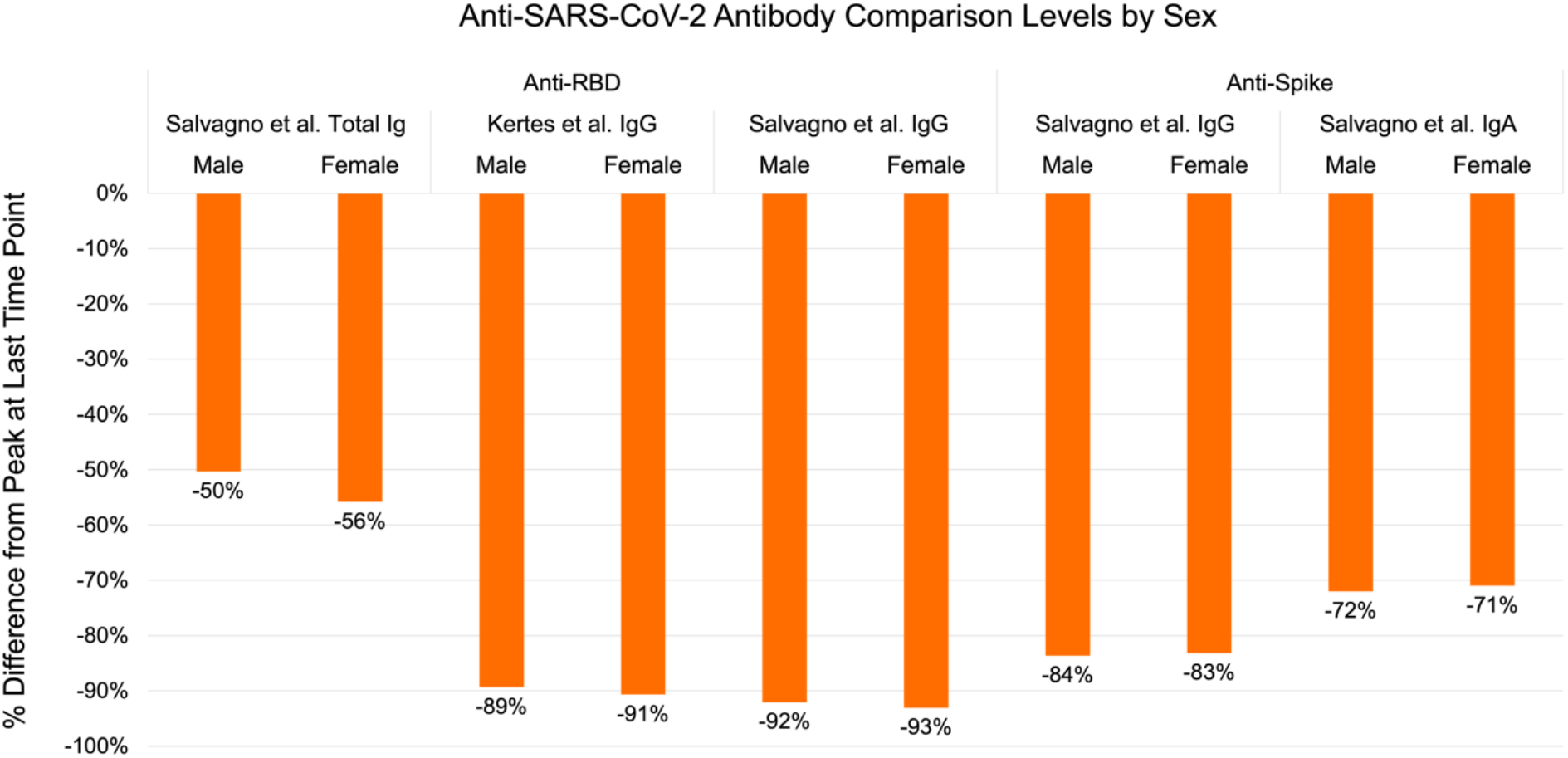
Comparison by sex of the percentage decline of anti-SARS-CoV-2 antibodies from peak to last measured timepoint (∼ 6 months) in each study by immunoglobulin class and molecular target.

**Figure 5.**
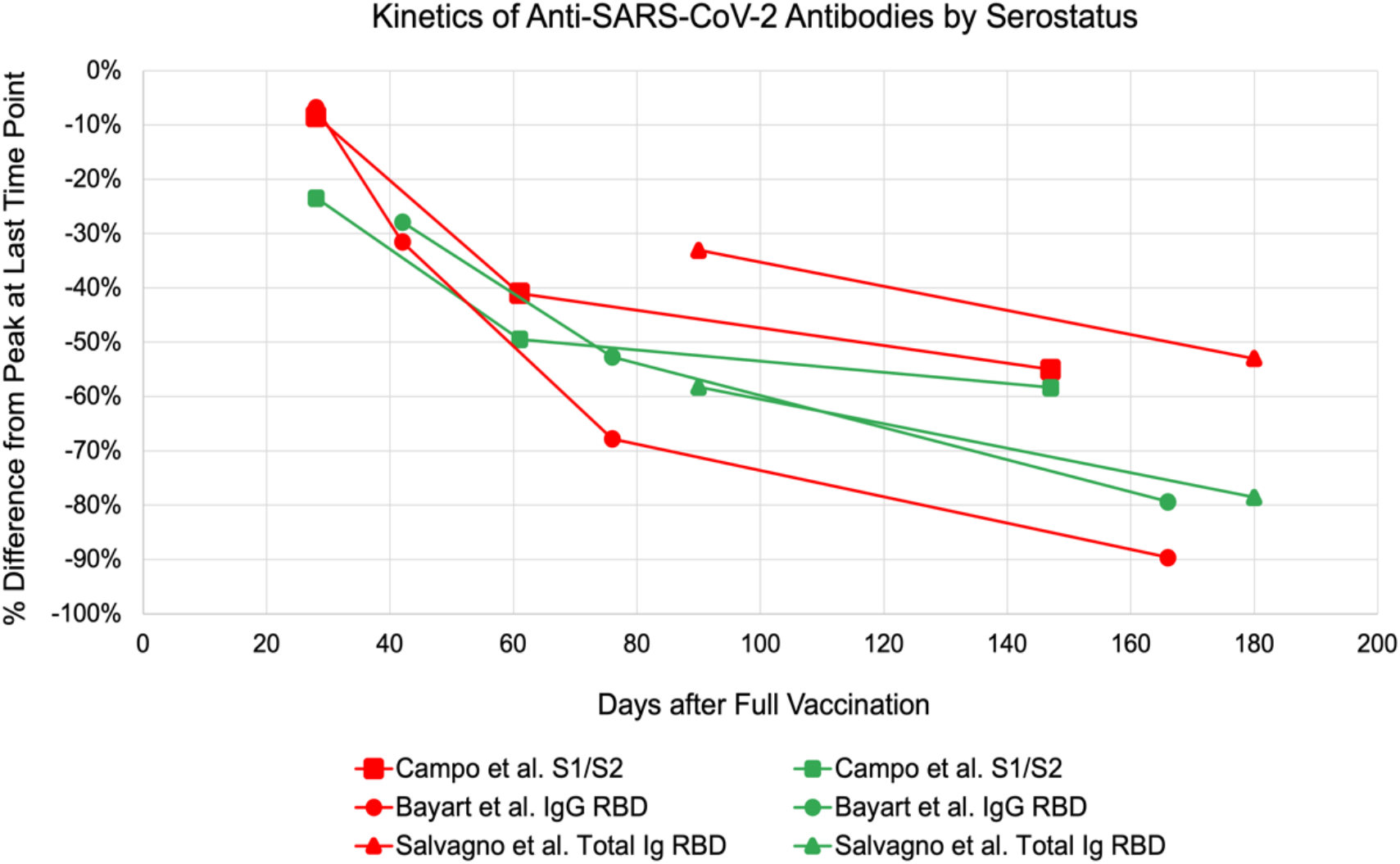
Comparison of antibody levels of seronegative and seropositive kinetics in studies reporting longitudinal data showing percentage decline from peak titer after vaccination. Red lines indicate seronegative data and green lines indicate seropositive data.

Although most of the studies focused on IgG titers, one study measured the levels of anti-spike IgA with a decline up to 72% at 150 days after its peak, consistent with the rate of decay of other immunoglobulins. Conversely, a study from Italy reported the decline in total Ig reaching only up to 56% at 150 days post-peak, which is lower than the rate of decline measured in other studies.

## Discussion

### Declining Ab levels post-mRNA vaccination

In the midst of the COVID-19 pandemic, the effectiveness and lasting protective effects of anti-SARS-CoV-2 vaccines are topics of interest for the general population. Here, we highlight the trends of Ab titers post-mRNA vaccination (**Table 1** and **Figure 2**). Most studies reported Ab peak titers within 1-4 weeks post-second dose vaccination, followed by a steady decline up to 90% in anti-SARS-CoV-2 Abs by 4-6 months post-vaccination. Consequently, as weeks post-mRNA vaccination increased, some studies reported an increase in the proportion of vaccine recipients whose Ab titers were below estimated protective thresholds.^17, 18^ Some studies noted that at 6 months post-vaccination, 16.1% of the study population had non-protective levels of anti-RBD IgG.^17^ Similarly, other studies reported that at 6 months post-vaccination, 95% and 27% of the study population had anti-RBD IgG and anti-spike trimeric IgG levels below the protective threshold, respectively.^18^ Although results are slightly dependent on the immunoassay used, the kinetics of Ab titer decline display similar properties.^18^

Across all studies, the rate of decline was noted to be greater during the initial weeks after reaching peak levels compared to 4-6 months post-mRNA vaccination (**Figure 2**). While the Ab level needed for protection is not established yet, there is a correlation between seroimmunity and protection from infection as well as seroimmunity and the likelihood of transmission.^19^ Therefore, this declining trend, with data demonstrating up to 95% decline in Ab levels depending on the antigenic target, supports the need for boosters and serologic monitoring moving forward to identify low responders and timely need for additional booster administration.

This trend was also observed in both Pfizer-BioNTech and Moderna mRNA vaccines, and regardless of the vaccine brand used, the absolute Ab titers of those with older age, seronegative status, or presence of major comorbidities were consistently lower at each time point, findings consistent with previous studies.^29^ Finally, with respect to the percentual decline in Ab titers 6 months post-second dose mRNA vaccination, there is a consistent rate of declining humoral response regardless of age, sex, serostatus, and comorbidities. Possible effects of the aforementioned factors on declining humoral response post-second dose mRNA vaccination are summarized in **Tables 2-5** and discussed in subsequent sections.

### Contributing factors to declining humoral immunity following complete mRNA vaccination Age

Both humoral and cellular immune responses are impaired as individuals age, ultimately leading to poorer vaccine response.^30^ Studies looking into the relationship of age and humoral response after undergoing Pfizer-BioNTech vaccination noted an inverse relationship on the antibody titers mounted.^12, 31^ The studies included in **Table 2** highlighted a similar trend. Regardless of the age group, there was a decline of titers starting one month after the second dose of vaccination (**Figure 3**). The mounted antibody titers are initially higher in the age group <60 years, but the rate of decline was consistent across all age groups. These all suggest the need for a booster vaccine dose for all age groups, but in low resource settings, prioritization should be given to older age groups who mount weaker humoral responses and overall lower antibody titers

### Sex

Ab titers of females were higher even after humoral decline in all studies (**Table 3**); however, the percentage decline from the peak was similar for both sexes (**Figure 4**).^12, 25^ While it is important to note that overall Ab titers may be influenced by the larger female population cohort (6,238 females *vs.* 3,128 males) involved in the included studies, this might be attributed to the sex-based difference in humoral immunity which affects vaccine responses. Previous studies indicated that females have a greater humoral response attributed to hormonal differences and X chromosomes leading to biallelic expression of certain genes.^33^ Several studies support this given the higher Ab titers mounted by females after vaccination against SARS-CoV-2.^31, 34^ Further, studies on the morbidity/mortality relationship due to SARS-CoV-2 infection between sexes found that males have three times higher probability of requiring intensive care unit admission and higher odds of mortality.^35^ While these studies point to the increased risk and mortality among males, the overall percentage decline of Ab titers identified post-second dose vaccination indicates the need for boosters regardless of sex.

### Serostatus

Infection with SARS-CoV-2 elicits a humoral immune response *via* production of immunoglobulins targeting the spike, nucleocapsid, as well as other viral proteins.^36^ This translates in detectable Ab titers prior to SARS-CoV-2 mRNA vaccine administration among previously infected individuals, and significantly higher Ab levels after the first dose compared to those with no history of prior infection.^36^ However, Ab titers measured after the second vaccine dose report varying trends (**Figure 5**). Some studies reported that seropositive groups continue to have increased Ab levels compared to seronegative groups, while others reported that there is no significant difference between the two groups.^37–42^ These disparities in trend are conveyed in the rate of decline of Ab titers at 5-6 months after full immunization compared to peak levels. Among the four journal articles included in this study, two reported that seropositive groups had a larger percent decline in Ab titers from peak compared with seronegative groups.^10, 12^ This is possibly due to higher peak values making Ab catabolism more pronounced in seropositive groups.^12^ Another possible explanation is due to timing, as seropositive groups reach peak Ab titers right after the first dose. Studies have observed that a second vaccine dose in seropositive groups does not significantly boost antibody titers higher, therefore these groups start their decline from peak much earlier than seronegative groups, who reach peak levels only after the second dose.^36, 38^ Conversely, two studies reported that seronegative groups had a larger percent decline.^43, 44^ This led to a recommendation to prioritize seronegative groups for booster administration over seropositive in cases of limited booster supply.^44^ However, given the heterogeneity of these findings, as well as a poor understanding of the immunity arising from the combination of native infection supplemented by vaccinated, booster administration should likely be recommended to everyone regardless of serostatus.

### Comorbidities

Several studies established that elderly patients (age >60 years) with stage-specific comorbidities along with other factors such as sex and serostatus elicit a comparably diminished humoral response post-vaccination.^29, 45^ Moreover, the robustness of initial Ab response post-vaccination is reported to predict rapidity of Ab titer decline.^19^ Those with higher maximum Ab response within 2 months from full vaccination were less likely to become seronegative or develop SARS-CoV-2 infection.^19^ As such, it is important to follow the course of Ab titers several weeks after full vaccination among those with stage-specific comorbidities in order to predict better when best to give additional booster vaccine doses and ensure appropriate protection.

In this systematic review, all three studies involving populations with comorbidities reported a consistent decrease in Ab titers from peak values at 4 months or later post-second dose SARS-CoV-2 mRNA vaccination.^19, 25, 46^ Interestingly, studies reported that the percent decline of Ab titer for those with chronic kidney disease, heart disease, diabetes mellitus, and immunosuppressive disorders was greater at 120 days after peak levels (28-50 days after first dose) compared to at 150 days after.^25^ These findings, however, may be attributed to the study design, as it was a non-serial study where the sample size at day 120 was less than at day 150, which could have skewed the results. This particular study was also limited by the lack of control groups, preventing us from directly comparing the percent decline of those with comorbidities *versus* healthy populations, although the absolute values of Ab levels were significantly lower in those with comorbidities compared to the total sample population.^25^ Regardless of this limitation, when taken together with other studies in this systematic review, those with comorbidities exhibited a similar trend in percent decline with the general population and other cohorts.

## Conclusion

In this systematic review, we characterized the substantial decline in Abs at ∼6 months after COVID-19 mRNA vaccinations. To date, several studies identified a significant correlation between time-dependent waning of Ab levels and an increased risk for breakthrough infections.^47, 48^ In addition to the rise of SARS-CoV-2 VoC, like the Delta and more recently the Omicron variant, studies also suggest a correlation between cold weather and an increase in COVID-19 cases.^49^ This is consistent with the established association between the circulation of respiratory viruses and climatic factors, displaying a peak incidence in the winter months.^49^ As such, the waning immunity, new VoC such as Omicron, uncontrolled community transmission of the Delta variant, and winter seasons could mark the culmination of major waves of COVID-19, which could in part be abated by booster doses. Indeed, boosters are likely to provide cross-reactive protection against SARS-CoV-2 infection, including VoC.^50^

Individuals in the older age group and with pre-existing comorbidities have lower levels of Abs compared to other individuals.^29^ Hence, even with the same rate of decline in humoral immunity post-second dose mRNA vaccination as observed in our study, their titers are consistently lower, and may in many cases be below adequate thresholds for protection. Thus, it is prudent to prioritize the elderly and those with comorbidities for booster administration in the coming months.

As evidence of waning Ab levels accumulate, several countries have expanded their vaccine coverage to include giving off booster doses to all adults after completion of primary vaccination.^51, 52^ As such, the rate of confirmed infection and the rate of severe illness were lower by a factor of 11.3 and 19.5 respectively, in the group that received the booster vaccine.^53^ A secondary analysis had also shown that the rate of confirmed infection at least 12 days after booster administration was lower than the rate after 4 to 6 days by a factor of 5.4.^53^ Further research is needed to understand the kinetics of antibody responses after vaccine boosters and to determine if a similar rate of decline is seen following adjunctive doses as observed with initial vaccination regimen.

In conclusion, our findings show that the levels of protective Abs significantly and consistently decline 6-8 months after the second dose of anti-SARS-CoV-2 mRNA vaccines regardless of age, sex, baseline serostatus and comorbidities. This supports that the rate of Ab decline is mostly independent from patient-related factors and peak Ab titer achieved after the second mRNA dose, but instead mainly a function of time and Ab class/molecular target, with some variability arising from the sensitivity of the immunoassay.^54^ Given this information, it may then be possible to predict the decline in Ab levels over time to an insufficient titer, based only on the patient’s peak Ab titer (∼4 weeks after second dose for seronegative patients), and thus identify in advance the optimal time when a booster vaccine dose should be indicated without the need for serial serologic monitoring. Such a strategy should be investigated for those who are likely to be low responders or at high risk of breakthrough infection.^55^ A similar strategy could be used following booster vaccine doses upon evaluation of Ab kinetics. Alternatively, research should explore the use of age, sex, serostatus, and comorbidity to gauge the most suitable time to measure the antibody titer in order to assess the need for a booster dose based on the data presented in our report. Given the significant decline of Abs observed in our systematic review, as well as studies showing correlation between Ab titers and breakthrough infections, it is of utmost importance to provide equal opportunities in obtaining the vaccines, create more efficient vaccination strategies, and to emphasize the necessity of providing booster shots in attenuating the effects of waning immunity, especially in the appearance of new VoC, in addition to the forthcoming winter season.

## Data Availability

All data produced in the present work are contained in the manuscript.

## Credit Authorship Contribution Statement

**Kin Israel Notarte:** Conceptualization, Visualization, Methodology, Validation, Formal analysis, Data curation, Writing – original draft, Writing – review & editing. **Israel Guerrero-Arguero:** Conceptualization, Formal analysis, Data curation, Writing – review & editing. **Jacqueline Veronica Velasco:** Methodology, Validation, Formal analysis, Data curation, Writing – original draft, Writing – review & editing. **Abbygail Therese Ver:** Methodology, Validation, Formal analysis, Data curation, Writing – original draft, Writing – review & editing. **Maria Helena Santos de Oliveira:** Formal analysis, Data curation, Writing – review & editing. **Jesus Alfonso Catahay**: Methodology, Validation, Formal analysis, Data curation, Writing – original draft, Writing – review & editing. **Md. Siddiqur Rahman Khan:** Writing – review & editing. **Adriel Pastrana**: Methodology, Validation, Formal analysis, Data curation, Writing – original draft, Writing – review & editing. **Grzegorz Juszczyk:** Writing – review & editing. **Jordi B. Torrelles:** Writing – review & editing. **Giuseppe Lippi:** Writing – review & editing. **Luis Martinez-Sobrido:** Writing – review & editing. **Brandon Michael Henry:** Conceptualization, Visualization, Validation, Formal analysis, Writing – review & editing.

## Ethical Approval

Not required.

